# Reporting of Fairness Metrics in Clinical Risk Prediction Models: A Call for Change

**DOI:** 10.1101/2024.03.16.24304390

**Authors:** Lillian Rountree, Yi-Ting Lin, Chuyu Liu, Maxwell Salvatore, Andrew Admon, Brahmajee K Nallamothu, Karandeep Singh, Anirban Basu, Bhramar Mukherjee

**Affiliations:** Department of Epidemiology, School of Public Health, University of Michigan, Ann Arbor, Michigan; Division of Pulmonary and Critical Care Medicine, Department of Internal Medicine, University of Michigan; Department of Epidemiology, School of Public Health, University of Michigan, Ann Arbor, Michigan; and VA Center for Clinical Management Research VA Ann Arbor Healthcare System, Ann Arbor; Institute for Healthcare Policy & Innovation, University of Michigan, and Veterans’ Affairs Center for Clinical Management Research, University of Michigan, and Michigan Center for Health Analytics and Medical Prediction, Department of Internal Medicine, University of Michigan Medical School, Ann Arbor; Department of Medicine, University of California San Diego School of Medicine, San Diego, California; The Comparative Health Outcomes, Policy, and Economics (CHOICE) Institute, School of Pharmacy, the Department of Health Systems and Population Health, and the Department of Economics, University of Washington, Seattle; Department of Biostatistics, School of Public Health, University of Michigan, Department of Epidemiology, School of Public Health, University of Michigan Ann Arbor, Michigan, USA

## Abstract

Clinical risk prediction models integrated in digitized healthcare systems hold promise for personalized primary prevention and care. Fairness metrics are important tools for evaluating potential disparities across sensitive features in the field of prediction modeling. In this paper, we seek to assess the uptake of fairness metrics in clinical risk prediction modeling by conducting a scoping literature review of recent high impact publications in the areas of cardiovascular disease and COVID-19. Our review shows that fairness metrics have rarely been used in clinical risk prediction modeling despite their ability to identify inequality and flag potential discrimination. We also find that the data used in clinical risk prediction models remain largely demographically homogeneous, demonstrating an urgent need for collecting and using data from diverse populations. To address these issues, we suggest specific strategies for increasing the use of fairness metrics while developing clinical risk prediction models.

## Introduction and Background

Prediction models are increasingly prevalent in research and decision making across a wide swath of fields, from finance and criminal justice to public health and healthcare (1,2). However, potential statistical and historical biases ingrained in these models can impact their accuracy and ethical viability, particularly for populations at risk of discrimination due to sensitive features like race/ethnicity, age, and sex (1,3–5). Are these seemingly objective and data-driven prediction models furthering existing inequities? As prediction models often inform who receives an intervention (e.g., a loan, a release on bail, a healthcare treatment) or identify who is at a higher disease risk when designing targeted prevention strategies, it is critical that they are fair, providing both accurate and nondiscriminatory predictions.

Algorithmic fairness is closely related to but distinct from algorithmic bias, another concern when assessing model performance (6). Algorithmic bias refers to the systematic and unfair discrimination that can occur when algorithmic models perpetuate existing biases present in the data they are trained on or the way they are designed. This bias can manifest in various ways, such as favoring one group over another due to race/ethnicity, sex, age, or other sensitive characteristics, or reinforcing stereotypes present in the training data. Algorithmic fairness, on the other hand, is the goal of designing algorithms and artificial intelligence models in a way that minimizes or mitigates bias and ensures fair treatment for all individuals or groups affected by the model (2,4,6,7). Achieving algorithmic fairness often requires careful consideration of the design, development, and deployment of models, including the selection of appropriate training data, the use of fairness-aware models, and the incorporation of fairness metrics to evaluate the performance of the model.

In recent years, several metrics have been introduced to evaluate the fairness of prediction models (1,8), alongside various packages and toolboxes for their implementation (see (9–12)). These metrics differ from common prediction metrics of discrimination and calibration, which measure overall model performance (6). Some of the most cited and used fairness metrics are enumerated in Table 1, all of which assess differences in predictions (*Ŷ*) for a binary decision outcome *Y* (e.g., treatment or no treatment, disease, or no disease) for different values of a sensitive variable *S* (*S=a* or *S=b*). These metrics give a numerical sense of the differences in predicted model outcomes across different demographic groups such as race/ethnicity, age, and sex, and thus how fair or unfair a model may be. The common practice is to use these metrics to provide only point estimates without associated uncertainty quantifications; while fair inferential methods have been proposed (13), statistical inference and interval estimation is not yet widely adopted in fairness research.

**Table 1.**
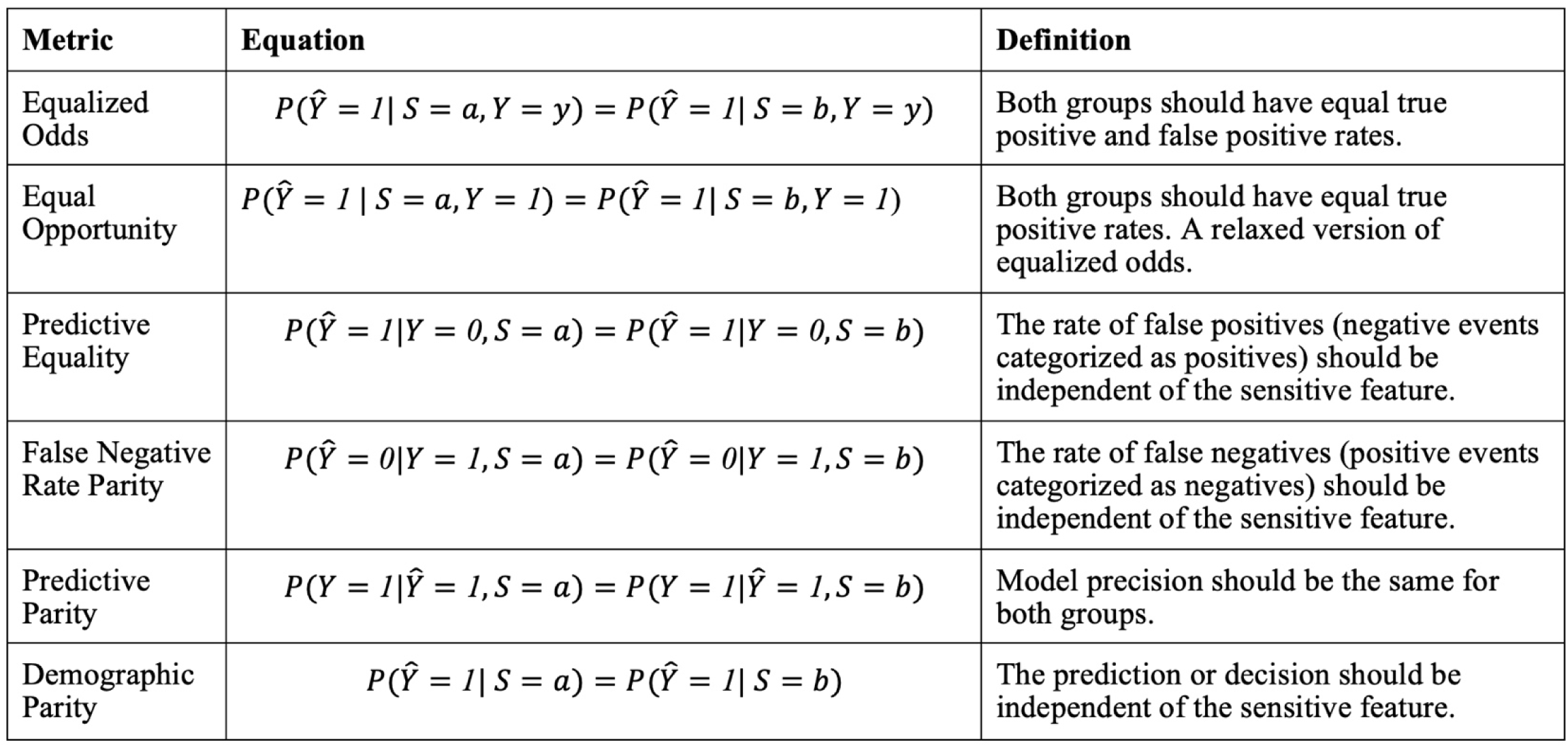
Commonly used fairness metrics, adapted from (1,8). Y-hat is our prediction/decision, Y is our observed data, and *S* is our observed feature, in the case of *S* being a multi-group variable where a comparison with a reference or privileged group is meaningful. Y-hat and Y are binary variables.

Though quite simple, such metrics can shed light on otherwise unseen disparities in a model or source dataset. No metrics, however, are without their limitations or challenges. For these metrics to produce meaningful results, a clear interpretation of the predictor variables used in a model is required in addition to how they affect the outcome of interest. For some variables, this is straightforward. For the sensitive features that fairness metrics usually seek to assess, however, it can be much more complex. Sensitive features often have multiple definitions and interpretations, particularly when it comes to what is biological versus what is socially constructed. This ambiguity is particularly important for sex and race/ethnicity, two of the most used variables in outcome regression models. Sex is a biological variable, but it will likely include impacts caused by individual’s gender—information that might be the actual cause behind the sex variable’s recorded influence on an outcome of interest. In the case of race and ethnicity, these variables’ self-reported—and thus exclusively social nature—is not always made clear, allowing for the inaccurate and harmful conclusion that differences observed with these race and/or ethnicity variables have a biological basis. Table 2 offers definitions of different measures related to these two specific sensitive variables and highlights how interpretations can be conflated and may not align with intended use.

**Table 2.**
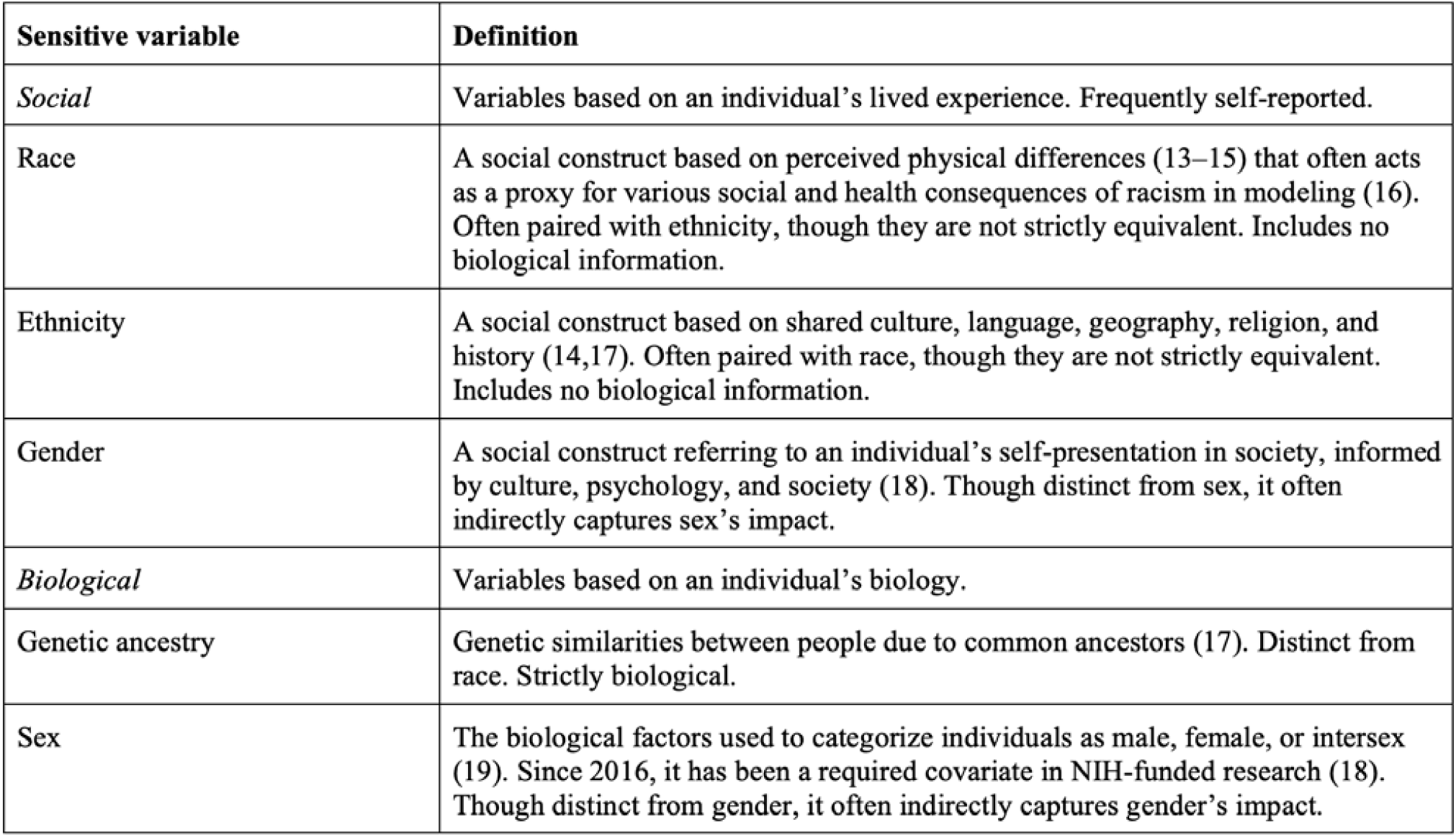
Definitions of commonly used sensitive variables. Note that the definition of these variables is frequently left ambiguous in practice, raising the possibility for harmfully conflating the social and the biological.

The risk of including sensitive variables only to misinterpret them in a way that furthers existing inequities has led to an ongoing debate about the place of these sensitive features in clinical disease risk prediction models. Exclusion of such sensitive features is one possible solution (one that has been particularly brought up regarding self-reported race (21,22)); the use of fairness metrics is another, allowing for the quantification of possible harm or help a sensitive variable might bring within the model. As prediction models are increasingly embedded into electronic health records and become more and more important for precision medicine (23,24), determining the place of sensitive features in prediction modeling is a crucial issue. Debates around best practice are ongoing, particularly regarding the inclusion of a race variable. Many other excellent papers explore the complexities of the issue at length (see (1,3,4,6,17,25)).

While assessing fairness metrics may be a key mechanism to ensure that models are equitable, how widely they are used or reported in the clinical risk prediction literature is not always objectively quantified. Thus, we sought to examine the usage of fairness metrics in clinical risk prediction research through a scoping review of recently published risk prediction models in high-impact journals for two diseases: cardiovascular disease (“CVD,” a long-studied chronic and non-communicable disease) and COVID-19 (a newly emerged infectious disease). We hypothesized that there would be little reporting of fairness metrics in CVD research, where many studies span years and began long before discussions of fairness metrics, but that the emergence of COVID-19 would pose an opportunity to more frequently incorporate modern advances in fairness metrics into predicting disease outcomes.

## Methods

A literature review was conducted for each of the two diseases of interest, CVD and COVID-19. Our outcomes of interest differed slightly between CVD and COVID-19: the clinical risk prediction models for CVD focused on fatal and non-fatal risk of CVD, while the models for COVID-19 focused on both risk of mortality and risk of severe disease (see Figure 1 for specific search terms). We did not use a classic systematic literature review approach, as our priority was to expediently capture only the highest impact papers. Figure 1 details the reproducible steps taken for data collection. This review began with searches on PubMed conducted in November 2023 to gain a high-level understanding of the use of sensitive features in risk prediction for CVD and COVID-19. Google Scholar was then used for actual article collection, focusing on extensively cited publications in high impact journals.

**Figure 1.**
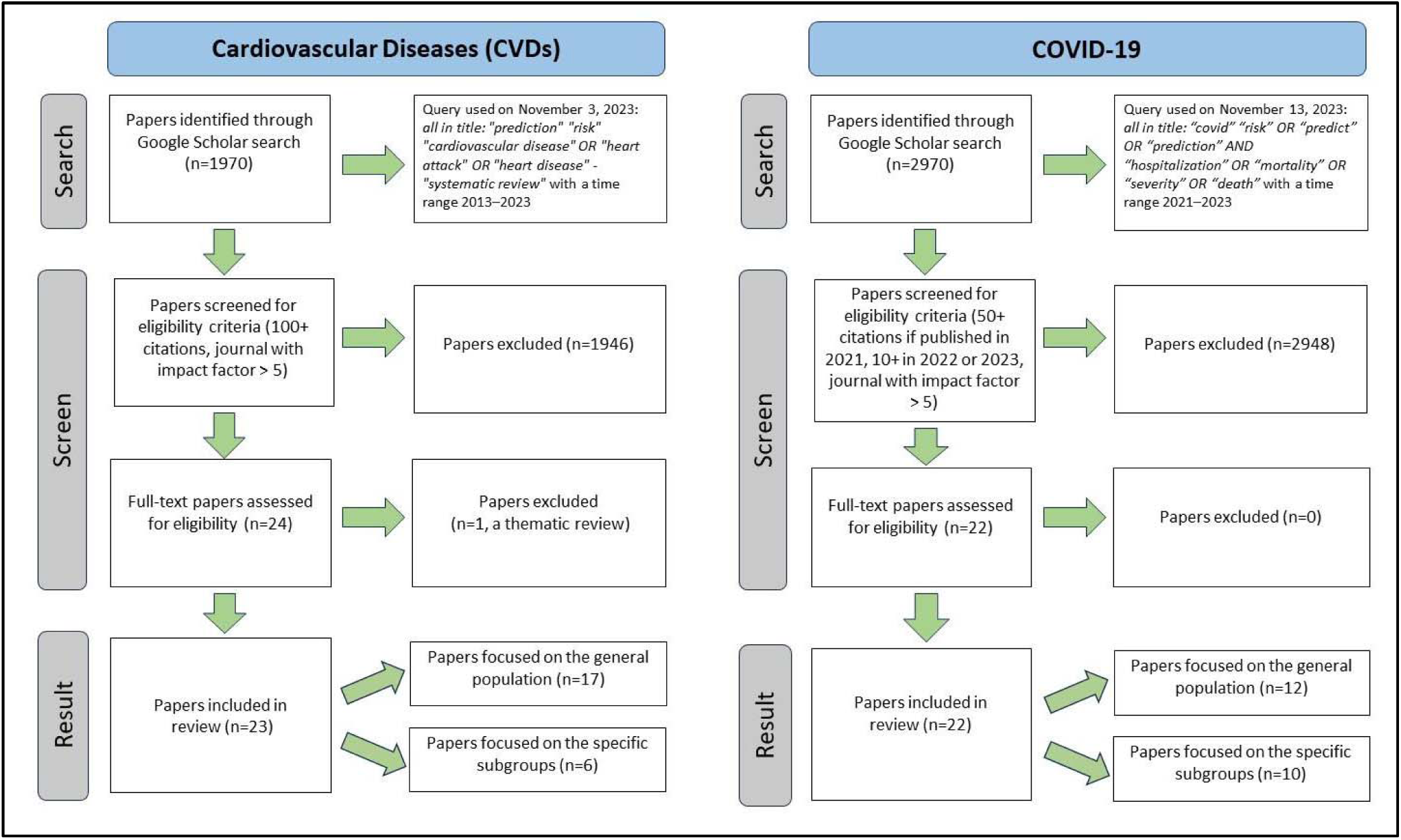
Flow diagram of the literature search process.

The criteria for highly impactful publications differed between CVD and COVID-19, reflecting the difference between a long-studied disease and an emerging area of study: CVD papers from the last ten years (2013–2023) were reviewed and selected if they exceeded 100 citations and were from journals with impact factors exceeding 5, while COVID-19 papers from 2021 to 2023 were reviewed and selected if they exceeded 50 citations for 2021 papers or exceeded 10 citations for 2022 to 2023 papers, both from journals with impact factors over 5. Systematic reviews and meta-analysis papers were excluded from the search results. Figure 1 is a flow diagram representing the search process.

## Results

### CVD

A PubMed search query *(“cardiovascular disease”[All Fields] OR “heart disease”[All Fields] OR “heart attack”[All Fields]) AND “prediction”[All Fields] AND “risk”[All Fields] AND (y_10[Filter]*) conducted on November 13, 2023, returned 5,107 results. Further specification with the term “sex” returned 817 results (16% of papers). The addition of the term “race” (but not “sex”) returned 145 results (2.8% of papers); specifying only one race/ethnicity returned 145 results (“Black”), 56 results (“Hispanic”), and 186 results (“Asian”). The Google Scholar search query: *allintitle: “prediction” “risk” “cardiovascular disease” OR “heart attack” OR “heart disease” OR “mortality” OR “death” -“systematic review”*, with a time range of 2013– 2023 on November 3, 2023, returned 1970 results, 1000 of which were accessible for review. The yielded results were selected if they <u>exceeded 100 citations</u> and were from journals with <u>impact factors exceeding 5</u>. This provided a shortlist of 23 articles detailing models predicting the risk of a fatal or non-fatal CVD event. These 23 articles were then divided into groups based on their target population (general population or a specific subpopulation). Sections S.1 and S.2 of the supplementary material provide additional details.

Of the 17 CVD papers focusing on a general population that met the criteria of this review (<u>supplementary material section S.1</u>), none discussed fairness metrics. Of these 17 papers, five (29%) stratified their models by sex, (i.e., built different models for each sex), and 11 (65%) included sex as a covariate. Nine of the 17 (53%) included data on race/ethnicity. As many of the papers paired race and ethnicity together or used them interchangeably, we will refer to any discussion of either as race/ethnicity jointly, despite this being an imprecise practice. Seven papers included race/ethnicity by self-reporting, and two by genetic ancestry (in genetically homogeneous populations). The eight studies that did not include race/ethnicity data were all based in the United States and Northern or Western Europe. Of the nine papers with recorded race/ethnicity data, five (55%) were multiracial/multiethnic (more than one racial/ethnic group identified). Four of the five multiracial/multiethnic studies included race/ethnicity as a covariate; no study stratified its model by race/ethnicity (see Figure 2). Other sensitive features considered in the studies include a covariate for area-based measures of deprivation (26), a covariate for body mass index (27), and stratification by risk region, a grouping of countries in Europe by their CVD mortality rates (28).

**Figure 2.**
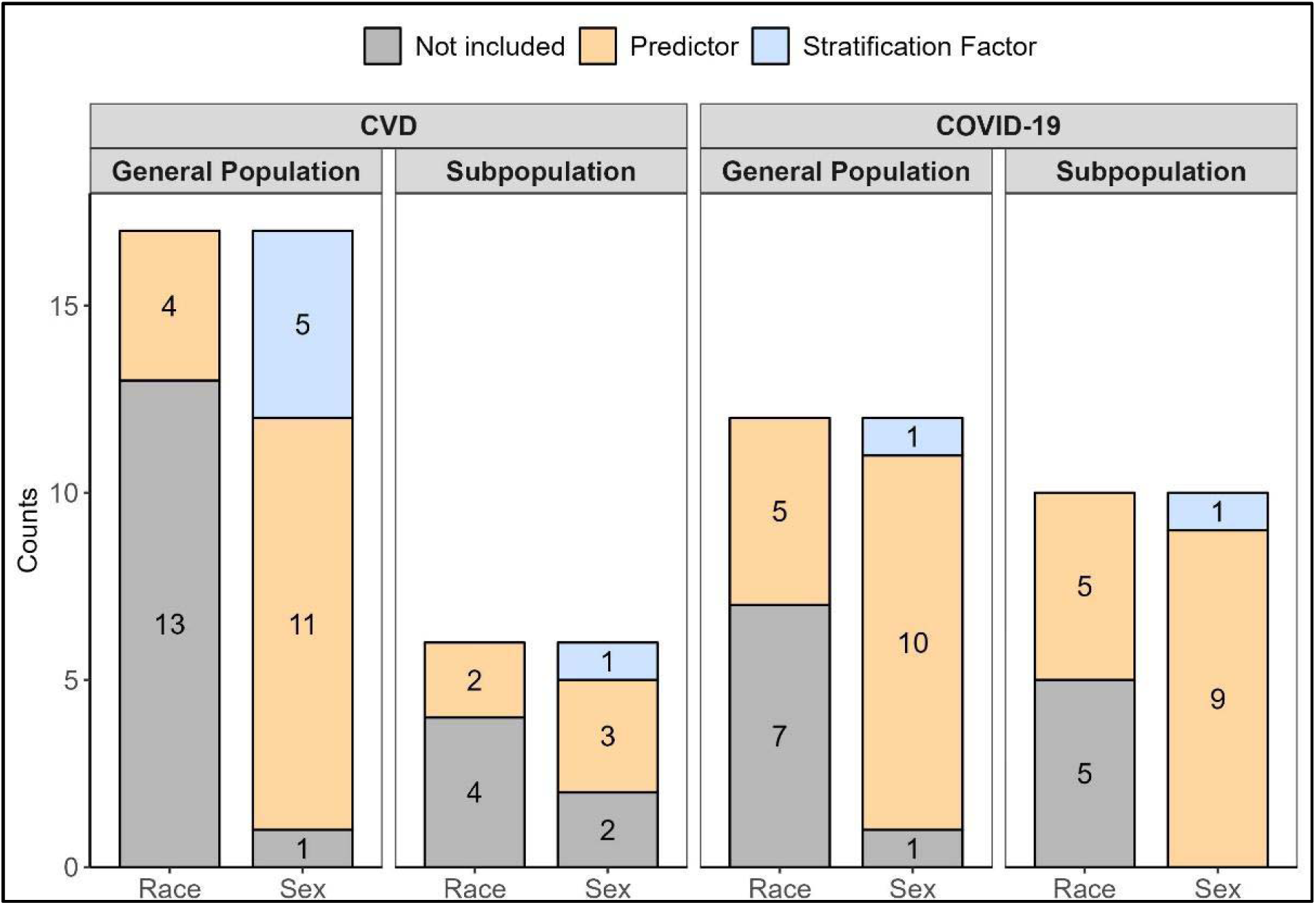
Counts depicting how many of the reviewed articles include race/ethnicity and/or sex as either predictors or stratification in their clinical risk prediction models, separated by disease of interest and population focus of the article. For cardiovascular diseases (CVD) the outcome considered was risk of cardiovascular disease, heart attack or heart failure, whereas for COVID-19 it was hospitalization and death.

Similar results were observed in the six CVD papers that focused on specific subpopulations (<u>supplementary material section S.2</u>). Subpopulations considered in these studies range from those with chronic conditions (29–32) to those from specific ethnic (33) or age groups (34). Three of the six studies (50%) included sex as a covariate; only one stratified its model by sex (see Figure 2). All but one study (34) included race/ethnicity data (all self-reported), but only three (50%) were multiracial/multiethnic, and only two (33%) consider race/ethnicity as a risk factor. Other sensitive features considered in the studies include covariates for geographic region and level of urbanization (33) and covariates measuring obesity (30).

### COVID-19

For COVID-19, the PubMed search query for all fields *(((covid) AND (risk)) AND (prediction) AND (2021/1/1:2023/12/31[pdat])) AND ((((hospitalization) OR (death)) OR (severity)) OR (mortality) AND (2021/1/1:2023/12/31[pdat])) NOT (((long) AND (meta)) AND (review) AND (2021/1/1:2023/12/31[pdat]))* conducted on November 13, 2023, returned 5722 results. The addition of the term “race” returned 141 results (2.46% of papers). The addition of the term “sex” (but not “race”) returned 614 results (10.73% of papers). Specifying only one race/ethnicity returned 103 results (“Black”), 62 results (“Hispanic”), and 71 results (“Asian”). The Google Scholar search query: *allintitle: “covid” “risk” “predict” OR “prediction” AND “hospitalization” OR “mortality” OR “severity” OR “death”* with a time range of 2021–2023 on November 13, 2023, returned 2970 results, 1000 of which were accessible for review. These results were then selected if they <u>exceeded 50 citations of 2021</u> papers and <u>exceeded 10 citations</u> <u>of 2022 to 2023 papers</u>, both from journals with impact factors over 5. This yielded a shortlist of 22 articles detailing models predicting the risk of COVID-19 hospitalization or death. These 22 were then divided into groups based on their target population (general population or a specific subpopulation). Sections S.3 and S.4 of the supplementary material provide details.

Of the 12 COVID-19 papers (<u>supplementary material section S.3</u>) focusing on a general population that met the inclusion criteria, none mention fairness metrics. Most papers (10 out of 12 papers, 83.33%) have sex as a covariate included in the prediction model; only one paper (8.33%) stratified by sex. Five out of 12 papers (41.67%) considered race/ethnicity, all five of which included it as a covariate—none stratified by race/ethnicity (see Figure 2). Of the remaining seven papers, three cited a lack of diverse data as the reason for excluding race/ethnicity information; one assumed the population to be entirely white (35); and the other three made no comments on race/ethnicity. Other potentially sensitive covariates explored include patient-level socioeconomic index (35) and the patient’s geographical region or hospital region (4 out of the 12 papers).

Similar results were observed in the list of ten COVID-19 papers (<u>supplementary material</u> <u>section S.4</u>) that focused on specific subpopulations. Subpopulations considered in the list range from those with pre-existing chronic diseases (36,37) to papers focused on the elderly (38) and infants (39). All ten papers included age as a risk factor; similarly, all ten included sex in their models. Only one paper implemented stratification by sex, with the remaining nine out of 10 (90%) including sex as a covariate. In all five papers (50%) that include race/ethnicity data, the information is used as a covariate rather than a means of stratification. Though there was very little stratification by sex and none by race/ethnicity (see Figure 2), stratification was undertaken for various other risk factors, such as the type of medication used (40) and geographical location (41).

## Discussion and Conclusion

We found that while the practice of assessing differences in model performance for sensitive features like sex and race/ethnicity was common, the use of fairness metrics to evaluate clinical risk prediction models was rare. Even though COVID-19 model constructions began well after the field of fairness metrics was first developed, fairness metric usage and similar considerations of sensitive features were as absent among COVID-19 as they were among CVD studies. Though some studies (7 of 23 of the CVD papers, 9 of 22 of the COVID-19 ones) used various discrimination and calibration metrics to assess model fit across different subgroups (such as across race/ethnicity), such calibration analysis was not routinely carried out; regardless, high discrimination and calibration alone do not guarantee model fairness (42,43). It is clear that, as the field of algorithmic fairness has grown and other disciplines have begun integrating fairness metrics into their own predictive modeling (44–48), clinical risk prediction models have not kept up with the progress.

Our reviews suggest that one major reason slowing the uptake of fairness metrics in clinical risk prediction models is the lack of data from diverse populations, particularly in a racial/ethnic and geographical sense. Though over half of the studies in the identified high impact CVD papers were multiracial/multiethnic, the data used in these studies was still over 50% one race/ethnicity. While there was more racially/ethnically diverse data in the COVID-19 papers—of the ten papers with multiracial/multiethnic data, only five (50%) were still a majority one race/ethnicity—a lack of geographical diversity remained (see Figure 3). Most studies (22 out of the 23 CVD papers, 15 out of 22 of the COVID-19 ones) were from the Global North. Though important work has been done to improve clinical risk prediction by focusing on data from underrepresented subgroups, such as with African Americans in the Jackson Heart Study (49), the results from our review suggest that such equity-focused research remains far from the norm in this field. Our search was limited to Google Scholar’s database and could have missed important articles, but high-level PubMed search queries support this conclusion: the search query of *“fairness”[All Fields] AND “risk prediction”[All Fields]* on November 16, 2023 returned only 15 papers total, none of which met our criteria for inclusion in our review.

**Figure 3.**
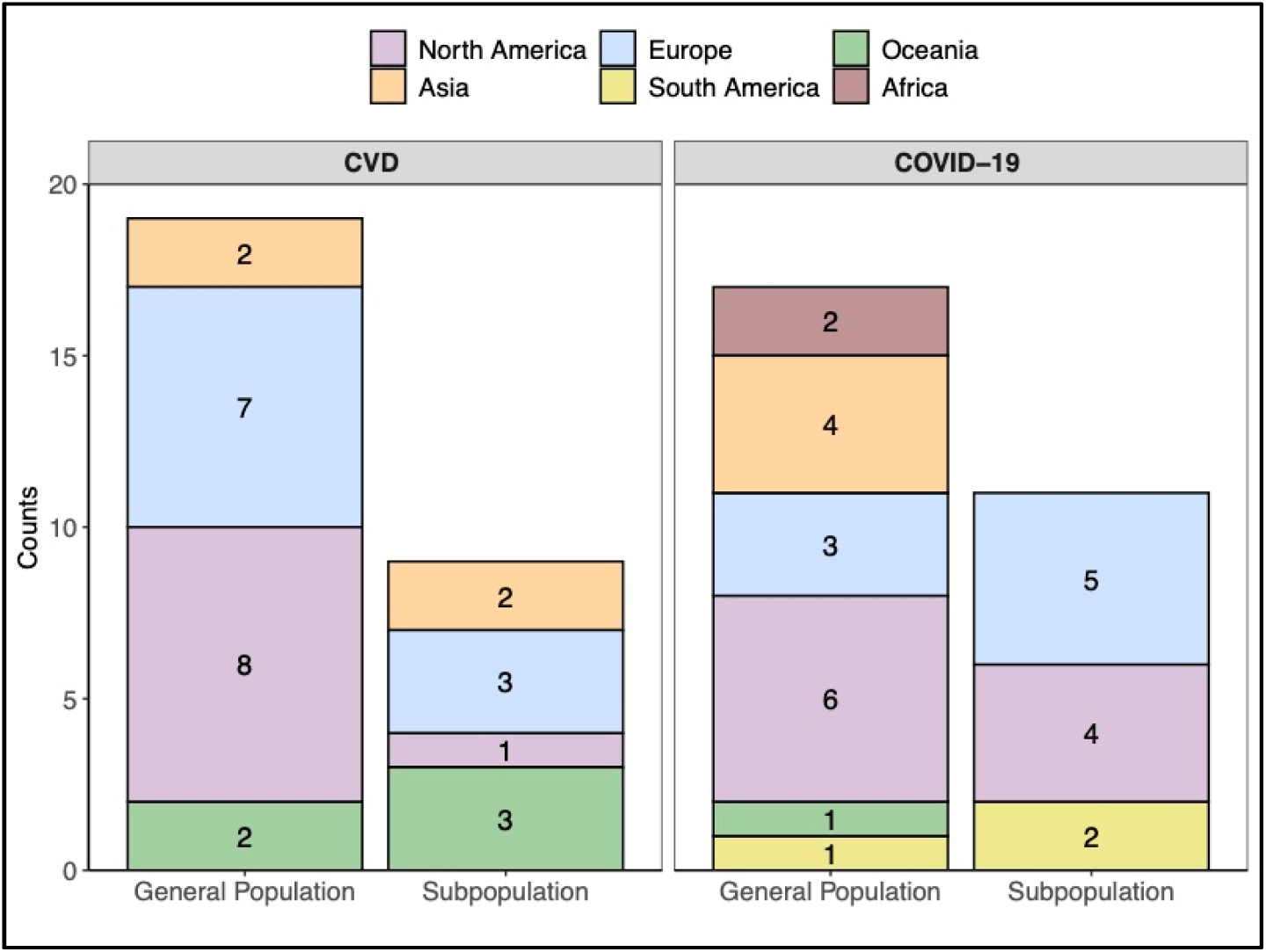
Counts depicting the geographic origin of data used in the reviewed articles, separated by disease of interest and population focus of the article. Studies with study regions covering multiple continents are double counted.

The largely homogenous data observed in our review implies that, for many studies, fairness assessment is infeasible to begin with. Lacking sufficient data to include meaningful covariates for different sensitive features, the inclusion of fairness metrics in analyses will be absent by default. Yet even when there clearly was the opportunity, such as the articles whose models were stratified by sex or study region, fairness analyses were not done. The opportunity to use fairness metrics is there; it simply has not been adopted as a part of the assessment routine.

It is possible that a lack of clarity as to how to best approach model fairness contributed to the dearth of fairness considerations seen in our review, as even when a clear opportunity for fairness metrics arises, the question of how to properly leverage them remains complex. There is no “one size fits all” method or universal fairness metric: instead, the specific context of the model—whether it is a preventative intervention or a limited-supply treatment, for example— must inform how relevant concerns of fairness are, and what metrics can address the concerns (6,7). Many fairness criteria are in fact mutually incompatible in practical settings (for example, demographic parity and equalized odds (2)), requiring a case-by-case decision on what kind of fairness (and thus fairness metric) will be most meaningful for the data and situation at hand. There is also the matter of the limitations of many of the most common fairness metrics that make their use inapplicable or unappealing for certain models. For example, most metrics are designed to assess only dichotomous outcomes and would require recalculation if a model’s predictions involved different cutoffs of the underlying continuous measures for different decisions. The “polarity” of a predicted outcome (where a “polar” outcome is one that is always preferred (6)) also impacts the importance of fairness, and identifying said polarity is not always straightforward (6,7). These limitations could be further contributing to the slow uptake in a clinical setting.

No critical number of developed fairness metrics will address this problem—it is not an issue of lack of methods, but of implementation. The methods exist and have already been adopted in a variety of other predictive modeling fields, including criminal justice, finance, and computational linguistics (46,48,50). There are signs of progress in clinical risk prediction, on both the applied (42) and theoretical (51) sides, but more work like these papers is needed. Our own recommendations for how this can happen in the field of clinical prediction are listed below and illustrated in the roadmap of Figure 4.

**Figure 4.**
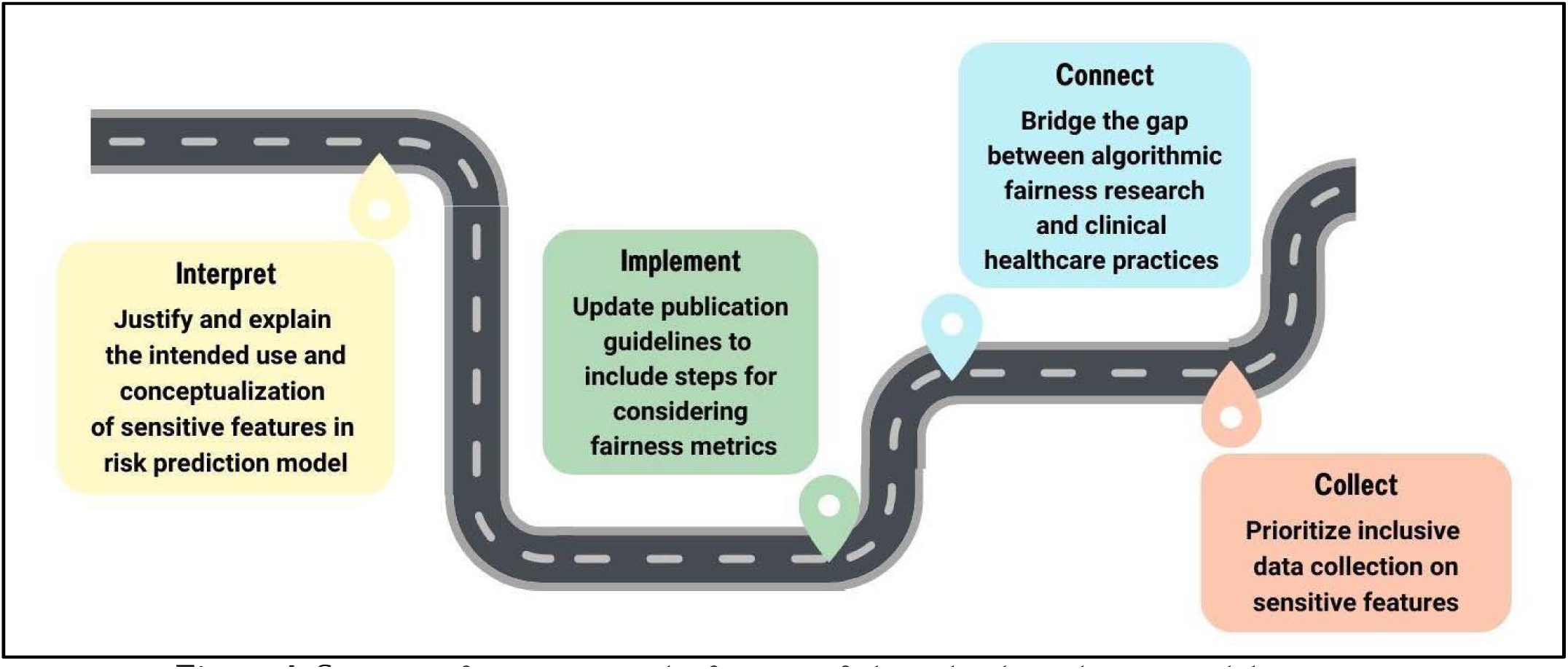
Strategies for increasing the fairness of clinical risk prediction models: Interpret, Implement, Connect and Collect (I2C2).

### Strategies for increasing the fairness of clinical risk prediction models across sensitive variables (I2C2)

● **<u>Interpret:</u>** In line with the NIH’s requirement of including (or justifying the exclusion) of a specifically biological sex variable, papers should interpret, justify, and explain their intended use and conceptualization of sensitive features in risk prediction models.
● **<u>Implement:</u>** Influential guidelines like EQUATOR’s TRIPOD guidelines (47, 48) should include steps on considering algorithmic fairness as part of implementation and application of clinical risk prediction models.
● **<u>Connect:</u>** The community of methods research in algorithmic fairness should ensure that the methods and tools developed are well broadcast to those in the community of practice in clinical healthcare, connecting theory to practice.
● **<u>Collect:</u>** The field of clinical risk prediction should highly prioritize collecting inclusive data across race, geographic region, and a variety of other sensitive or historically underrepresented features.

To understand the current barriers in the practice community, we have developed a short questionnaire for the authors of our selected studies or those like them to help the field identify key challenges in implementing fairness metrics. This questionnaire will help elucidate where resources should be most concentrated for this I2C2 roadmap. It can be <u>accessed here</u>, and Table 3 below shows the summary questions. We hope by educating and enabling practitioners around use of fairness metrics, we will create a more equitable prediction world for all.

**Table 3.** The ten questions included in the questionnaire. All but questions 9 and 10 are multiple choice, with the option to elaborate in a free response.

| **A 10-question survey for understanding investigators' barriers for using fairness metrics in clinical risk prediction models** |  |
| --- | --- |
| Question 1 | What is the overall goal of the clinical risk prediction model that you are constructing? |
| Question 2 | What were the sensitive features included in your prediction model? |
| Question 3 | What is the intended use of the sensitive features in your prediction models? |
| Question 4 | Are the measures (and how they were collected) consistent with their intended use? |
| Question 5 | How were the sensitive features you selected in Q2 included in your model? |
| Question 6 | What were the criteria used for model evaluation and performance? |
| Question 7 | Were the model evaluation criteria focused on overall performance or the performance within specific subgroups of the data defined by the sensitive variables you chose in Q2? |
| Question 8 | Was model fairness considered and/or assessed? |
| Question 9 | If you answered yes to Q7, please explain how you considered model fairness. If you answered no, please explain what prevented such consideration. |
| Question 10 | If there were any other challenges or insights concerning the use (or non-use) of model fairness in your developed prediction model that you would like to share, please do so below. |

All materials used for this review can be accessed via GitHub.

## Data Availability

All materials used for this review can be accessed via GitHub.

https://github.com/umich-cphds/fairness-metrics/

## Notes

### Competing Interest Statement

The authors have declared no competing interest.

### Funding Statement

This work was supported through grant DMS1712933 from the National Science Foundation and MI-CARES grant 1UG3CA267907 from the National Cancer Institute. The funders had no role in the design of the study; collection, analysis, or interpretation of the data; writing of the report; or the decision to submit the manuscript for publication.

