## Supplemental materials for "Reporting of Fairness Metrics in Clinical Risk Prediction Models: A Call for Change"

### Appendix
### S.1 Table of Selected CVD Papers for General Population

| Title | Authors | Link/DOI | Citations | Journal | Year | Outcome assessed | Type of model used | Geographic region | Racial demographics (self-reported unless otherwise noted) | Sensitive features considered as risk factors | Sensitive features considered for stratified analysis | Were model calibration and discrimination assessed for different sensitive features? | Criteria for model evaluation | Do authors explicitly consider or report fairness metrics? |
|---|---|---|---|---|---|---|---|---|---|---|---|---|---|---|
| Development and validation of QRISK3 risk prediction algorithms to estimate future risk of cardiovascular disease: prospective cohort study | Hippisley-Cox et al | 10.1136/bmj.j2099 | 923 | BMJ | 2017 | Risk of CVD | Cox's proportional hazards models | United Kingdom | Self-reported in ~60% of all subjects. Majority (>85%) White or not recorded. ~2% Indian. ~1% Pakistani. ~1% Bangladeshi. ~1.3% Other Asian. ~1% Black Caribbean. ~2% Black African. ~0.8% Chinese. 2.5% Other. | Age, ethnic origin | Sex | Yes | Harrell's C-statistics | No |
| Risk prediction of cardiovascular death based on the QTc interval: evaluating age and gender differences in a large primary care population | Nielsen et al | 10.1093/eurheartj/ehu081 | 123 | European Heart Journal | 2014 | Risk of CVD | Cox regression models | Denmark | Not included | Age | Sex | Yes | C-statistics, Brier scores | No |
| Cardiovascular disease risk prediction equations in 400 000 primary care patients in New Zealand: a derivation and validation study | Pylypchuk et al | 10.1016/S0140-6736(18)30664-0 | 224 | The Lancet | 2018 | Risk of fatal or non-fatal CVD event | Cox proportional hazard model | New Zealand | Majority (>55%) European. ~13% Maori. ~13% Pacific. ~9% Indian. ~11% Chinese or other Asian. | Measures of deprivation, ethnicity, age | Sex | Yes | $R^2$, Harrell's C statistic, and Royston's D statistic | No |
| The ACC/AHA 2013 pooled cohort equations compared to a Korean Risk Prediction Model for atherosclerotic cardiovascular disease | Jung et al | 10.1016/j.atherosclerosis.2015. 07.033 | 111 | Atherosclerosis | 2015 | Ten-year atherosclerotic cardiovascular disease risk | Cox proportional hazard models | South Korea | Korean | Age | Sex | Yes | C-statistic, Hosmer-Lemeshow | No |
| SCORE2 risk prediction algorithms: new models to estimate 10-year risk of cardiovascular disease in Europe | SCORE2 working group and ESC Cardiovascular risk collaboration | 1 0.1093/eurheartj/ehab309 | 397 | European Heart Journal | 2021 | Ten-year risk of fatal and non-fatal CVD | Fine and Gray model stratified by cohort and fitted to sex | Europe and North America | Not included | Age | Risk region, sex | Yes | Harrell's C-Index | No |
| Risk prediction by genetic risk scores for coronary heart disease is independent of self-reported family history | Tada et al | 10.1093/eurheartj/ehv462 | 276 | European Heart Journal | 2016 | Time to first occurrence of CHD | Cox proportional hazards regression models | Sweden | Swedish ancestry (genetic) | Sex, age | N/A | No | Wald tests | No |
| A Validated Model for Sudden Cardiac Death Risk Prediction in Pediatric Hypertrophic Cardiomyopathy | Miron et al | 10.1161/CIRCULATIONAHA.120.047235 | 121 | Circulation | 2020 | Risk of sudden cardiac death | Cause-specific hazard regression model | Canada, United States, Australia | Not included | Sex, age | N/A | No | C-statistic, cross validation | No |
| Multilocus Genetic Risk Scores for Coronary Heart Disease Prediction | Ganna et al | 10.1161/ATVBAHA.113.301218 | 174 | Arteriosclerosis, Thrombosis, and Vascular Biology | 2013 | Risk of coronary heart disease | Cox proportional hazard model | Sweden | Not included | Sex, age | N/A | No | C-index | No |

| Title | Authors | Link/DOI | Citations | Journal | Year | Outcome assessed | Type of model used | Geographic region | Racial demographics (self-reported unless otherwise noted) | Sensitive features considered as risk factors | Sensitive features considered for stratified analysis | Were model calibration and discrimination assessed for different sensitive features? | Criteria for model evaluation | Do authors explicitly consider or report fairness metrics? |
|---|---|---|---|---|---|---|---|---|---|---|---|---|---|---|
| Improving the accuracy of prediction of heart disease risk based on ensemble classification techniques | Latha and Jeeva | 10.1016/j.imu.2019.100203 | 435 | Informatics in Medicine Unlocked | 2019 | Risk of heart disease | Comparitive analysis of various classification ML algorithms | United States | Not included | Sex, age | N/A | No | Ten-fold cross validation | No |
| Comparison of machine learning algorithms for clinical event prediction (risk of coronary heart disease) | Beunza et al | 10.1016/j.jbi.2019.103257 | 142 | Journal of Biomedical Informatics | 2019 | Coronary risk at ten years | Machine learning classification algorithms | United States | Not included | Sex, age | N/A | No | AUC | No |
| An improved ensemble learning approach for the prediction of heart disease risk | Mienye et al | 10.1016/j.imu.2020.100402 | 136 | Informatics in Medicine Unlocked | 2020 | Risk of CVD event | Ensemble learning | United States | Not included | Sex, age | N/A | No | ROC | No |
| Validation of the 2014 European Society of Cardiology Guidelines Risk Prediction Model for the Primary Prevention of Sudden Cardiac Death in Hypertrophic Cardiomyopathy | Vriesendorp et al | 1 0.1161/CIRCEP.114.002553 | 143 | Circulation: Arrhythmia and Electrophysiology | 2015 | Five-year risk of sudden cardiac death | Cox regression models | Belgium and the Netherlands | Not included | Age | N/A | No | ROC, C-statistics | No |
| Endothelial Dysfunction, Increased Arterial Stiffness, and Cardiovascular Risk Prediction in Patients With Coronary Artery Disease: FMD-J (Flow-Mediated Dilation Japan) Study A | Maruhashi et al | 10.1161/JAHA.118.008588 | 106 | Journal of the American Heart Association | 2018 | Risk of recurrent cardiovascular events | Cox proportional hazard regression analysis | Japan | Japanese | Sex, age | N/A | No | Schoenfeld residuals | No |
| Electrical risk score beyond the left ventricular ejection fraction: prediction of sudden cardiac death in the Oregon Sudden Unexpected Death Study and the Atherosclerosis Risk in Communities Study | Aro et al | 10.1093/eurheartj/ehx331 | 115 | European Heart Journal | 2017 | Sudden cardiac arrest | Multivariable logistic regression analysis | United States | In cases: 82% white, 11% Black, 1.9% Hispanic, 5.2% other. In controls: 92% white, 3.5% Black, 1.4% Hispanic, 3.3% other. | Sex, age | N/A | No | Hosmer-Lemeshow, C-statistic | No |
| Genetic Risk Prediction and a 2-Stage Risk Screening Strategy for Coronary Heart Disease | Tikkanen et al | 1 0.1161/ATVBAHA.112.301120 | 206 | Arterioscleros is, Thrombosis, and Vascular Biology | 2013 | Fatal and non-fatal CHD risk | Cox regression models | Finland | Finnish ancestry (genetic) | Sex, age | N/A | No | C-index | No |
| 10-Year Coronary Heart Disease Risk Prediction Using Coronary Artery Calcium and Traditional Risk Factors: Derivation in the MESA (Multi-Ethnic Study of Atherosclerosis) With Validation in the HNR (Heinz Nixdorf Recall) Study and the DHS (Dallas Heart Study) | McClelland et al | 10.1016/j.jacc.2015.08.035 | 561 | Journal of the American College of Cardiology | 2015 | Incident CHD events | Penalized Cox proportional hazards model | United States | A combination of several datasets. One was entirely (100%) Caucasian. The largest was 38.5% Caucasian, 11.8% Chinese American, 27.8% African American, 22% Hispanic American. The other was 37.9% Caucasian, 49.1% African American, 11.3% Hispanic American, 1.8% Other. | Age, sex, BMI, race/ethnicity | N/A | Yes | C-statistic and ROC | No |

| Title | Authors | Link/DOI | Citations | Journal | Year | Outcome assessed | Type of model used | Geographic region | Racial demographics (self-reported unless otherwise noted) | Sensitive features considered as risk factors | Sensitive features considered for stratified analysis | Were model calibration and discrimination assessedfor different sensitive features? | Criteria for model evaluation | Do authors explicitly consider or report fairness metrics? |
|---|---|---|---|---|---|---|---|---|---|---|---|---|---|---|
| Global Electric Heterogeneity Risk Score for Prediction of Sudden Cardiac Death in the General Population | Waks et al | 10.1161/CIRCULATIONAHA.116.021306 | 131 | Circulation: Arrhythmia and Electrophysiology | 2016 | Risk of sudden cardiac death | Cox proportional hazards models | United States | 77.3% white; 22.7% Black (other races excluded) | Sex, race, age | N/A | No | Schoenfeld residuals, C-statistics | No |

### S.2 Table of Selected CVD Papers for Subpopulation

| Title | Authors | Link/DOI | Citations | Journal | Year | Outcome assessed | Type of model used | Geographic region | Racial demographics (self-reported unless otherwise noted) | Sensitive features considered as risk factors | Sensitive features considered for stratified analysis | Were model calibration and discrimination assessed for different sensitive features? | Criteria for model evaluation | Do authors explicitly consider or report fairness metrics? |
|---|---|---|---|---|---|---|---|---|---|---|---|---|---|---|
| Predicting the 10-Year Risks of Atherosclerotic Cardiovascular Disease in Chinese Population | Yang et al | 10.1161/CIRCULATIONAHA.116.022367 | 402 | Circulation | 2016 | Ten year risk of fatal and non-fatal CVD | Cox proportional hazards models | China | Chinese | Age, geographic region (Northern/Southern China), urbanization | Sex | Yes | C-statistics and modified Nam-D'Agostino test | No |
| Cardiovascular Disease Risk Prediction in the HIV Outpatient Study | Thompson-Paul et al | 10.1093/cid/ciw615 | 146 | Clinical Infectious Diseases | 2016 | Risk of CVD event | Adaptations of Framingham, PCEs, SCORE and DAD | United States | Majority (>50%) White, non-Hispanic. ~30% Black, non-Hispanic. ~12% Hispanic. ~3% Other. | Sex, age | N/A | No | C-statistic and Hosmer-Lemeshow | No |
| An updated prediction model of the global risk of cardiovascular disease in HIV-positive persons: The Data-collection on Adverse Effects of Anti HIV Drugs (D:A:D) study | Friis-Møller et al | 10.1177/2047487315579291 | 228 | European Journal of Preventative Cardiology | 2020 | Risk of CVD event | Cox regression models | Europe and Australia | Majority (>60%) White. ~7% Non-White. 32% Unknown. | Age, sex, ethnicity | N/A | No | C-statistic and Hosmer-Lemeshow | No |
| Development of a Novel Risk Prediction Model for Sudden Cardiac Death in Childhood Hypertrophic Cardiomyopathy (HCM Risk-Kids) | Norrish et al | 10.1001/jamacardio.2019.2861 | 135 | JAMA Cardiology | 2019 | Risk of sudden cardiac death or equivalent event | Cox proportional hazards regression models | Western Europe, Eastern Europe, Japan, Australia | Not included | None | N/A | No | Schoenfeld residuals, C-index | No |
| Anthropometric measurements of general and central obesity and the prediction of cardiovascular disease risk in women: a cross-sectional study | Goh et al | 10.1136/bmjopen-2013-004138 | 204 | BMJ Open | 2014 | Ten-year CVD risk | Framingham risk score model, SCORE risk chart for high-risk regions, general CVD and simplified general CVD risk score models | Australia | Reported as ethnicity: Australia (76.5%); UK and Ireland (9.5%); Northern Europe (4.1%); Southern Europe (5.4%); Asia (4.5%). | Obesity, ethnicity | N/A | No | ROC | No |
| Prediction of First Cardiovascular Disease Event in Type 1 Diabetes Mellitus | Vitisen et al | 10.1161/CIRCULATIONAHA.115.018844 | 104 | Circulation | 2016 | Fatal and non-fatal CVD risk | Poisson regression analysis | Denmark | White (>90% Danish ancestry) | Sex, age | N/A | No | Hosmer-Lemeshow | No |

### S.3 Table of Selected COVID-19 Papers for General Population

| Title | Authors | Link/DOI | Citations | Journal | Year | Outcome assessed | Type of model used | Geographic region | Racial demographics (self-reported unless otherwise noted) | Sensitive features considered as risk factors | Sensitive features considered for stratified analysis | Were model calibration and discrimination assessed for different sensitive features? | Criteria for model evaluation | Do authors explicitly consider or report fairness metrics? |
|---|---|---|---|---|---|---|---|---|---|---|---|---|---|---|
| Predicting mortality risk in patients with COVID-19 using machine learning to help medical decision-making | Pourhomayoun and Shakibi | 10.1016/j.smhl.2020.100178 | 246 | Smart Health | 2021 | Risk of mortality | Multiple ML methods | 146 countries | Not included | Age, sex, country, province, city, pre-existing health conditions | N/A | No | Accuracy, sensitivity, specificity, AUC | No |
| Clinical, radiological, and laboratory characteristics and risk factors for severity and mortality of 289 hospitalized COVID-19 patients | Zhang et al. | 10.1111/all.14496 | 204 | Allergy | 2021 | Risk of in-hospital mortality | Logistic regression | China | Not included | Age, sex, comorbidity, smoking, surgery history, biological measurements | N/A | No | AUC | No |
| Clinical and inflammatory features based machine learning model for fatal risk prediction of hospitalized COVID-19 patients: results from a retrospective cohort study | Guan et al. | 10.1080/07853890.2020.1868564 | 109 | Annals of Medicine | 2021 | Risk of mortality | XGBoost | China | Not included | Disease severity, age, biological measurements | N/A | No | AUC, prediction accuracy, precision, and F1 scores | No |
| Risk factors for COVID-19 progression and mortality in hospitalized patients without pre-existing comorbidities | Liu et al. | 10.1016/j.jiph.2021.11.012 | 26 | Journal of Infection and Public Health | 2022 | Risk of mortality | Logistic regression | China | Not included | Sex, age, biological measurements | N/A | No | Not specified | No |
| Machine learning approaches in Covid-19 severity risk prediction in Morocco | Laatifi et al. | 10.1186/s40537-021-00557-0 | 27 | Journal of Big Data | 2022 | Risk of severity | UMAP, logistic regression, SVM, KNN, GaussianNB, decision tree | Morocco | Not included | Sex, age, comorbidities, biological measurements | N/A | No | Accuracy, sensitivity, specificity, AUC | No |
| External validation of the QCovid risk prediction algorithm for risk of COVID-19 hospitalisation and mortality in adults: national validation cohort study in Scotland | Simpson et al. | 10.1136/thoraxjnl-2021-217580 | 17 | Thorax | 2022 | Risk of hospitalization, risk of death | QCOVID algorithm (Fine-Gray sub-distribution hazard model) | Scotland, United Kingdom | Not included | Socioeonomic status and pre-existing health conditions | Sex, age, admission period | No | Harrell's C, $R^2$, D-statistics, Brier Score | No |
| Predicting the evolution of COVID-19 mortality risk: A recurrent neural network approach | Villegas et al. | 10.1016/j.cmpbup.2022.100089 | 13 | Computer Methods and Programs in Biomedicine | 2023 | Risk of mortality | RNN | Spain | Not included | Sex, age, medication, hospital stay, biological measurement | N/A | No | Accuracy, sensitivity, specificity, AUC | No |
| 19–Related Hospitalization, Intensive Care Unit Admission, Invasive Mechanical Ventilation, and Death — United States, March–December 2020 | Kompaniyets et al. | 10.15585/mmwr.mm7010e4 | 394 | Morbidity and Mortality Weekly Report | 2021 | Risk of hospitalization, risk of ICU admission, risk of death | Multivariable logit model | United States | Hispanic (10.4 %), White non-Hispanic (63.7%), Black non-Hispanic (18.4%), Asian non-Hispanic (2.1%), Other (4%), Unknown (1.4%) | Sex, race, hospital region, insurance payer type | Age, BMI | No | Sensitivity analysis | No |

| Title | Authors | Link/DOI | Citations | Journal | Year | Outcome assessed | Type of model used | Geographic region | Racial demographics (self-reported unless otherwise noted) | Sensitive features considered as risk factors | Sensitive features considered for stratified analysis | Were model calibration and discrimination assessed for different sensitive features? | Criteria for model evaluation | Do authors explicitly consider or report fairness metrics? |
|---|---|---|---|---|---|---|---|---|---|---|---|---|---|---|
| Risk factors for mortality in patients with COVID-19 in New York City | Mikami et al. | 10.1007/s11606-020-05983-z | 286 | Journal of General Internal Medicine | 2021 | Risk of in-hospital mortality | Generalized additive models and Cox proportional hazard regression model | United States | White (26.9%), Black (24.1%), Asian (4.4%), Others (44.7%), Hispanic (25.4%), Non-Hispanic (57.5%), Unknown ethnicity (17%) | Age, sex, race, smoking, ethnicity, biological measurements, comorbidities | N/A | No | AUC | No |
| Federated learning of electronic health records to improve mortality prediction in hospitalized patients with COVID-19: machine learning approach | Vaid et al. | 10.2196/24207 | 100 | JMIR Medical Informatics | 2021 | Risk of mortality | Multilayer perceptron (MLP) model, logistic regression, federate learning model | United States | Hispanic (26.07%), non-Hispanic (59.45%), Unknown ethnicity (14.48%); White (23.9%), Black (28.54%), Asian (4.84%), Other (38.57%), Unknown (4.14%) | Sex, race, ethnicity, past medical history | N/A | No | AUC | No |
| Estimating risk of mechanical ventilation and in-hospital mortality among adult COVID-19 patients admitted to Mass General Brigham: The VICE and DICE scores | Nicholson et al. | 10.1016/j.eclinm.2021.100765 | 83 | EClinical Medicine | 2021 | Ventilation in COVID Estimator [VICE] score and Death in COVID Estimator [DICE] score | Logistic regression | United States | White (42%), Black (17.9%), Hispanic (10.8%), Asian (3.6%), Other/mix (17%), Not recorded (8.6%) | Weight, age, sex, BMI, comorbidities, race, biological measurements, and medication taken | N/A | No | AUC and C-statistics | No |
| Machine-learning-based COVID-19 mortality prediction model and identification of patients at low and high risk of dying | Banoei et al. | 10.1186/s13054-021-03749-5 | 63 | Critical Care | 2021 | Mortality outcome and clustering of high mortality risk patients | Statistically inspired modification of partial least square (SIMPLS) analysis | United States | European American (36.75%), African American (15.25%), Asian (1%), More than one race (5%), Hispanic (59.75%), non-Hispanic (40.25%) | Age, sex, race, smoking, alcohol, level of consciousness, mental status | N/A | No | $Q^2$, $R^2$, and AUC | No |

### S.4 Table of Selected COVID-19 Papers for Subpopulation

| Title | Authors | Link/DOI | Citations | Journal | Year | Outcome assessed | Type of model used | Geographic region | Racial demographics (self-reported unless otherwise noted) | Sensitive features considered as risk factors | Sensitive features considered for stratified analysis | Were model calibration and discrimination assessed for different sensitive features? | Criteria for model evaluation | Do authors explicitly consider or report fairness metrics? |
|---|---|---|---|---|---|---|---|---|---|---|---|---|---|---|
| Clinical characteristics and risk factors for death among hospitalised children and adolescents with COVID-19 in Brazil: an analysis of a nationwide database | Oliveira et al. | 10.1016/S2352-4642(21)00134-6 | 112 | The Lancet Child and Adolescent Health | 2021 | Time to recovery or time to death | Proportional sub-distribution hazards model | Brazil | White (35.24%), Black or Brown (62.42%), Asian (0.88%), Indigenous (1.44%) | Date of admission, date of onset, age, sex, ethnicity, geopolitical macroregion, number of health conditions | N/A | No | Competing risks analysis | No |
| Association between antidepressant use and reduced risk of intubation or death in hospitalized patients with COVID-19: results from an observational study | Hoertel et al. | 10.1038/s41380-021-01021-4 | 208 | Molecular Psychiatry | 2021 | Time from study baseline to intubation or death | Cox regression proportional hazard models | France | Not included | Sex, age, hospital, obesity, smoking status, medical condition, and biological markers of disease severity | Antidepressant type | No | Sensitivity analysis | No |
| Hypertension, diabetes and obesity, major risk factors for death in patients with COVID-19 in Mexico | Peña et al. | 10.1016/j.arcmed.2020.12.002 | 138 | Archives of Medical Research | 2021 | Risk of mortality | Logistic regression | Mexico | Not included | Age, sex, residence, smoking status, pneumonia, treat at home or hospital, and other chronic conditions | N/A | No | Not specified | No |
| Clinical characteristics and risk factors for mortality in very old patients hospitalized with COVID-19 in Spain | Ramos-Rincon et al. | 10.1093/gerona/glaa243 | 123 | The Journals of Gerontology: Series A | 2021 | Risk of in-hospital mortality | Logistic regression | Spain | Not included | Age, sex, comorbidities, dependence state, symptoms, biological measurements | N/A | No | Hosmer–Lemeshow test | No |
| Risk of infection, hospitalisation, and death up to 9 months after a second dose of COVID-19 vaccine: a retrospective, total population cohort study in Sweden | Nordström et al. | 10.1016/S0140-6736(22)00089-7 | 209 | The Lancet | 2022 | Risk of infection, risk of severe COVID | Cox regression proportional hazard models | Sweden | Not included | Age, sex, homemaker service, comorbidities, born in Sweden or not | N/A | No | Schoenfeld residuals | No |

| Title | Authors | Link/DOI | Citations | Journal | Year | Outcome assessed | Type of model used | Geographic region | Racial demographics (self-reported unless otherwise noted) | Sensitive features considered as risk factors | Sensitive features considered for stratified analysis | Were model calibration and discrimination assessed for different sensitive features? | Criteria for model evaluation | Do authors explicitly consider or report fairness metrics? |
|---|---|---|---|---|---|---|---|---|---|---|---|---|---|---|
| Risk prediction of covid-19 related death and hospital admission in adults after covid-19 vaccination: national prospective cohort study | Hippisley-Cox et al. | 10.1136/bmj.n2244 | 262 | BMJ | 2021 | Time to COVID-19 related death and time to hospitalization | Cause-specific Cox proportional hazard models | United Kingdom | White (68.77%), Indian (2.91%), Pakistani (1.61%), Bangladeshi (1.17%), Other Asian (1.68%), Caribbean (0.7%), Black African (1.63%), Chinese (0.6%), Other (2.7%) | Age, sex, ethnic origin, comorbidities, and dose of vaccination | N/A | No | C-statistics, Rsq, D statistics | No |
| Comparison of mortality risk in patients with cirrhosis and COVID-19 compared with patients with cirrhosis alone and COVID-19 alone: multicentre matched cohort | Bajaj et al. | 10.1136/gutjnl-2020-322118 | 207 | Gut | 2021 | Risk of mortality | Logistic regression | United States | White (56.25%), Non-white (43.75%), Hispanic ethnicity (7.35%), non-Hispanic ethnicity (92.65%) | Sex, age, race and ethnicity, smoking status, comorbid conditions | N/A | No | Not specified | No |
| Metformin and risk of mortality in patients hospitalised with COVID-19: a retrospective cohort analysis | Bramante et al. | 10.1016/S2666-7568(20)30033-7 | 156 | The Lancet Healthy Longevity | 2021 | Risk of in-hospital mortality | Logistic regression, mixed-effect logistic regression, Cox proportional hazard models, propensity-matched mixed-effects logistic regression | United States | Not included | Sex, age, pre-existing health condition, other medication use, and BMI | Metformin use and sex | No | Schoenfeld residuals, sensitivity analysis | No |
| Maternal vaccination and risk of hospitalization for Covid-19 among infants | Halasa et al. | 10.1056/NEJMoa2204399 | 112 | The New England Journal of Medicine | 2022 | Risk of hospitalization | Logistic regression | United States | White non-Hispanic (39.18%), Black non-Hispanic (17.54%), Hispanic any-race (28.79%), Other non-Hispanic(6.39%), Unknown (8.1%) | Age, sex, race and ethnicity, region | Type of vaccine, time of vaccine, and date of admission | No | Not specified | No |
| Diet quality and risk and severity of COVID-19: a prospective cohort study | Merino et al. | 10.1136/gutjnl-2021-325353 | 144 | Gut | 2021 | Risk of infection, risk of severe COVID | Cox regression proportional hazard models | United States and United Kingdom | White (96%), Black (0.7%), Asian (1.8%), Other (1.2%), Missing (0.3%) | health condition, zip code, social economic status, smoking, physical | Socio-economic status, dietary preference | No | Schoenfeld residuals | No |
